# Impact of SETD8/KMT5A Overexpression on Hepatocellular Carcinoma Progression and Prognosis

**DOI:** 10.1101/2025.11.11.25340025

**Authors:** Norihiko Suzaki, Shinya Hayami, Atsushi Miyamoto, Masashi Nakamura, Tomohiro Yoshimura, Kensuke Nakamura, Yoshinobu Shigekawa, Atsushi Shimizu, Yuji Kitahata, Akihiro Takeuchi, Hideki Motobayashi, Shogo Ehata, Ryuji Hamamoto, Manabu Kawai

## Abstract

**Background:** Hepatocellular carcinoma (HCC) has a poor prognosis due to its high recurrence rate, even after curative surgery. Epigenetic regulators play a critical role in cancer progression, and the histone methyltransferase SETD8/KMT5A has been reported to be overexpressed in various malignancies. This study aimed to elucidate the role of SETD8/KMT5A in HCC.

**Methods:** We investigated SETD8/KMT5A expression in 345 primary HCC resection specimens through immunohistochemical staining. For functional analyses, we conducted loss-of-function study of SETD8/KMT5A using HCC cell lines, including *in vitro* assays for proliferation, cell cycle, invasion, and RNA sequencing with gene ontology (GO) analysis. Additionally, we performed xenograft experiments in mice and performed similar experiments using the SETD8/KMT5A inhibitor UNC0379.

**Results:** All cases were divided into either the SETD8 high-expression (n=197) or low-expression (n=148) groups. The high-expression group exhibited significantly poorer 5-year overall survival and 2-/5-year disease-free survival compared with the low-expression group (both p<0.001). Multivariate analysis indicated that high SETD8 expression was an independent poor prognosis factor in overall (p=0.0255) and disease-free (p=0.0051) survival. SETD8/KMT5A knockdown suppressed proliferation by inhibition of G1 to S phase transition (p<0.001). GO terms were related to cancer progression, including cell adhesion, MAPK-related signaling and chromatin remodeling. SETD8/KMT5A knockout using CRISPR/Cas9 inhibited tumor growth (p<0.01) *in vivo*.

**Conclusions:** SETD8/KMT5A overexpression was associated with poor prognosis and was an independent prognostic factor in HCC. *In vitro* and *in vivo* analysis, the inhibition of SETD8 could repress HCC progression through the regulation of cell activity and cell cycle transition.

## Introduction

Hepatocellular carcinoma (HCC) is a malignancy that can lead to high rate of recurrence, even after curative resection.[1] In cases of unresectable or recurrent HCC, chemotherapy including multi-kinase inhibitors is currently secondary to immune checkpoint inhibitors,[2–5] but the effectiveness of these drugs has been limited. In HCC, molecular targeted drugs have thus far all relied on phosphorylation, so novel therapeutic agents targeting different pathways or mechanisms other than phosphorylation are desirable.[6] We previously reported the involvement of several histone methyltransferases/demethylases in poor prognosis for various malignancies, and we clarified further mechanisms for the progression of these tumors.[7–10] Regarding interactions between different post-translational modifications (PTMs), RB1 methylation by histone methyltransferase SMYD2 promoted RB1 phosphorylation and cell cycle progression.[11] Methylation might promote phosphorylation through post-translational modification interaction, but the detailed molecular mechanisms through methylation still require elucidation.

The target of the current study, SETD8 (also known as KMT5A or PR-SET7) is known as an enzyme that monomethylates lysine 20 on histone 4 (H4K20), and methylation for non-histone proteins such as p53 and PCNA has been reported.[12] SETD8 is reportedly associated with various malignancies, including bladder cancer, lung cancer, thyroid papillary carcinoma, ovarian cancer and neuroblastoma.[12–15] Based on these reports and our previous studies, we hypothesized that SETD8 might also be associated with HCC, which led us to select it as the focus of this study. Here, we therefore aim to elucidate the critical roles of SETD8/KMT5A by expression analysis using clinical HCC tissue samples, and loss-of-function analyses using SETD8-specific small interfering RNAs (siSETD8) *in vitro* and SETD8-specific clustered regularly interspaced short palindromic repeats (CRISPR)/CRISPR associated protein 9 (Cas9) system *in vivo*.

## Methods

This retrospective study was approved by the Wakayama Medical University Ethics Committee (approval number: 2521) and conducted in accordance with the principles of the Declaration of Helsinki. As this study used only existing clinical data and resected specimens, an opt-out approach was adopted, which is considered equivalent to informed consent under institutional guidelines. Information about the study was disclosed on the Wakayama Medical University website, allowing participants the opportunity to decline participation.

### Patients

This study focuses upon the 345 patients with HCC that underwent primary hepatectomy at our hospital between January 2000 and December 2014. Follow-up data were collected until 2019, due to allow the calculation of up to 5-year survival outcomes. Clinical data were accessed for research purposes in 2019. During data retrieval, authors were able to view identifiable patient information. However, all data were fully anonymized prior to analysis to ensure that individual patients could not be identified.

### Immunohistochemistry (IHC)

Slides with paraffin-embedded HCC specimens were deparaffinized with xylene and rehydrated with 100% ethanol. The slides were processed under high pressure (121℃ for 10 min) in Target Retrieval Solution Citrate pH9 (Dako, Glostrup, Denmark), and subsequently 0.3% hydrogen peroxide (H_2_O_2_). After being treated with Protein Block Serum-Free Ready-to-use (Dako), the slides were incubated overnight at 4°C with a mouse anti-SETD8 monoclonal antibody (ab3798: abcam, Cambridge, UK) at 1:2000 dilution ratio. The slides were reacted with anti-mouse EnVision HRP (Dako) and stained with Histofine DAB substrate kit (Nichirei, Tokyo, Japan) and Mayer’s hematoxylin solution (Fujifilm, Tokyo, Japan). IHC staining was evaluated with the immunoreactive scoring system (IRS).[16] Valuation was performed without knowledge of the clinical-pathological variables. The cases of HCC were divided into two groups based on the median IRS value as the cutoff, following previous studies: the SETD8 high and low expression groups. [17, 18]

### Cell culture

Experiments were conducted using two human HCC cell lines: SNU475 and HuH-7. HuH-7 cells were purchased from the Japanese Collection of Research Bioresources (JCRB) Cell Bank (Osaka, Japan), and SNU475 cells were obtained from the American Type Culture Collection (ATCC, Manassas, VA, USA). HuH-7 was cultured using Dulbecco’s modified Eagle’s medium (Thermo Fisher Scientific, Waltham, MA), while SNU475 was cultured using RPMI-1640 medium (Thermo Fisher Scientific).

### Transfection with small interfering RNA (siRNA)

Two types of siRNA targeting human SETD8 (siSETD8#1, siSETD8#2) and siRNA targeting enhanced green fluorescent protein (EGFP) as a negative control (siEGFP) were synthesized (Merck-Sigma-Aldrich, Darmstadt, Germany). The nucleotide sequences of the synthesized siRNAs were determined based on our previous research, and they are as shown in S1 Table. The transfection of siRNA into the HCC cell lines was performed using the Lipofectamine RNAiMAX method (Thermo Fisher Scientific) with a final concentration of 20nM.

### Quantitative Real-time Polymerase Chain Reaction (qRT-PCR)

Specific primers were designed for Glyceraldehyde-3-phosphate dehydrogenase (GAPDH, housekeeping gene), and SETD8. The nucleotide sequences of the primers were determined based on previous research and are as shown in S2 Table.[12] Transfected HCC cell lines with siSETD8 and siEGFP (final concentration: 20nM) were prepared, and total RNA was extracted using the RNeasy Mini Kit (QIAGEN, Venlo, Netherlands). Complementary DNA (cDNA) was synthesized from total RNA using SuperScript III RT (Thermo Fisher Scientific). The cDNA was combined with SsoAdvanced Universal SYBR Green Supermix (Bio-Rad Laboratories, Hercules, CA) and forward and reverse primers (50 nM final concentration), followed by qRT-PCR using CFX96 touch (Bio-Rad Laboratories). We analyzed the expression levels of messenger RNA and evaluated them using the ΔΔCt method.[19] The experiments were conducted in triplicate.

### Treatment with known SETD8-specific inhibitor: UNC0379

As it is thought to be close to the clinical application, we conducted experiments using UNC0379 trifluoroacetate salt (Merck-Sigma-Aldrich). Cells in culture were treated with SETD8-specific inhibitor UNC0379 at a concentration of 7.3 µM and incubated for 24 hours.[20] These cells were compared with cells to which PBS (the solvent for UNC0379) was added as a control.

### Cell proliferation assay

We seeded 96-well plates with HCC cells at a density of 1.0×10^4^ cells per well with pre-incubation for 24 hours. Subsequently, transfection with siRNA (final concentration: 20nM) or treatment with UNC0379 (7.3µM) was performed, followed by 24-hour incubation. Cell counting and evaluation of cell numbers were conducted using the Cell Counting Kit-8 (DOJINDO, Kumamoto, Japan). Absorbance was measured at 450 nm using a microplate reader, SH-9000 (Corona Electric, Ibaraki, Japan). The experiment was conducted in triplicate.

### Cell cycle analysis

The HCC cell lines were transfected with siRNA (final concentration: 20nM) or treated with UNC0379 (7.3µM) and incubated for 24 hours. Subsequently, the cells were processed according to the recommended protocol of the BrdU flow kit (BD Pharmingen, Franklin Lakes, NJ). We performed cell cycle analysis using the obtained cells with FACS Calibur (BD Pharmingen) and Cell Quest Pro software (BD Pharmingen). The experiment was conducted in triplicate.

### Cell invasion assay

HCC cell lines were transfected with siRNA (final concentration: 20nM) or treated with UNC0379 (7.3µM) and incubated for 24 hours. Subsequently, we evaluated cell invasion ability using the CytoSelect 24-well Cell Invasion Assay (CBA-110: Cell Biolabs, San Diego, CA) following the recommended protocol. The seeded cells were 2.0×10^5^. The experiment was conducted in triplicate.

### Differentially expressed genes (DEGs) and gene ontology (GO) analysis using RNA sequence

After treatment with siRNA (final concentration: 20nM), total RNA was extracted from HCC cells with RNeasy Mini kit (QIAGEN). We then requested RNA sequence analysis by Cancer Precision Medicine (Kanagawa, Japan). Gene Ontology analysis was performed with ClueGO (ver. 2.5.8) in Cytoscape (ver. 3.8.2). We compared RNA extracted from siSETD8-transfected cells with RNA extracted from siEGFP-transfected cells to identify differentially expressed genes (DEGs). GO terms and pathways were visualized using the Cluego software in Cytoscape, with consideration of the *p*-value obtained from the GO analysis.

### Transfection with clustered regularly interspace short palindromic repeats/ Cas9 (CRISPR/Cas9)

In preparation for conducting xenograft experiments in mice, it was necessary to generate permanent SETD8 knockout cells. We therefore conducted genome editing using the CRISPR/Cas9 system to create knockout cell lines. We also performed co-transfection with a Puromycin resistance gene to select the knockout cell lines. The plasmids used included Pr-Set7 CRISPR/Cas9 KO plasmid (h2) (sc-415759-KO-2, Santa Cruz Biotechnology, Dallas, TX), Pr-Set7 HDR plasmid (h2) (sc-41579-HDR-2, Santa Cruz Biotechnology), and control CRISPR/Cas9 plasmid (sc-418922, Santa Cruz Biotechnology). We performed plasmid transfection using UltraCruz Transfection Reagent (sc-395739, Santa Cruz Biotechnology) following the recommended protocol (plasmid DNA final concentration: 1µg/ml). Subsequently, 1 µg/ml puromycin (sc-108071, Santa Cruz Biotechnology) was added to the cell lines that underwent transfection treatment, and they were then incubated for 24 hours.

### Animal experiments

All animal procedures were performed based on the recommendations of the Guide for Care and Use of Laboratory Animals (National Research Council) and this study was approved by the Wakayama Medical University Animal Care and Use Committee (approval number: 1108). This study was conducted in accordance with the ARRIVE guidelines.

HCC cell lines of 5.0×10^6^ with SETD8 knockout and control knockout were inoculated into the subcutaneous right dorsal area of 6-week-old BALB/cSlc-nu/nu mice (Japan SLC, Shizuoka, Japan) using MatriMix (Nippi, Tokyo, Japan) as the substrate. The tumor sizes were measured every 7 days with calipers, and estimated tumor volumes were calculated with a formula:

Tumor volume = (length × width^2^) /2.

All mice were euthanized at 42 days after inoculation by cervical dislocation under isoflurane anesthesia. The experiment was conducted in triplicate.

### Statistical analysis

Statistical analyses were performed using JMP Pro 16 (SAS Institute, Cary, NC, USA) and Excel Statistics (BellCurve for Excel, version3.21; Social Survey Research Information Co., Ltd., Tokyo, Japan). Patient characteristics and laboratory data were analyzed using Fisher’s exact test and the Mann-Whitney U test. Overall survival curve and disease-free survival curve were analyzed by Kaplan-Meier method and log-rank test. Risk factors for overall survival and disease-free survival were analyzed using the Cox proportional hazards model. All experiments were performed in triplicate and Student’s t-test was used for statistical analysis. Statistical significance was defined as *p* < 0.05.

## Results

### Clinicopathological findings of SETD8 expression in HCC

Based on the immunohistochemical staining, patients with HCC were categorized into either the SETD8 high-expression group (n=197) with IRS of 6-12, or the SETD8 low-expression group (n=148) with IRS of 0-5. Representative IHC images corresponding to IRS are shown in Fig 1A. Patient characteristics such as sex, age, alcohol abuse and viral infection (hepatitis B virus or hepatitis C virus) are shown in Table 1; there was no statistical difference between the SETD8 high-expression and the low-expression groups. In preoperative laboratory findings, the SETD8 high-expression group had higher AFP levels (35.3 vs. 14.1 [ng/ml], *p*=0.008) and des-γ-carboxy prothrombin (DCP) levels (567 vs. 149 [mAU/ml], *p*<0.001) than the SETD8 low-expression group. In pathological findings, SETD8 high-expression group had larger tumor size, greater number of tumors, poorer differentiation and more vascular invasion than the SETD8 low-expression group (Table 1). In five-year overall survival after surgery, the SETD8 high-expression group had poorer prognosis than the low-expression group (Fig 1B, 47% vs 71%). In two- and five-year disease-free survival after surgery, the SETD8 high-expression group had poorer prognosis than the low-expression group (Fig 1C; 2-years: 39% vs 64%, 5-years: 26% vs 41%, respectively). In the multivariate analysis of overall survival, identified prognostic factors were: ICGR15 > 10%, AFP > 20 ng/ml, tumor size, TNM stage III-IV and high expression of SETD8. In the multivariate analysis of disease-free survival, recurrence risk factors were ICGR15>10%, AFP>20 ng/ml, multiple tumors, vascular invasion and high expression of SETD8 (Table 2).

**Fig 1.**
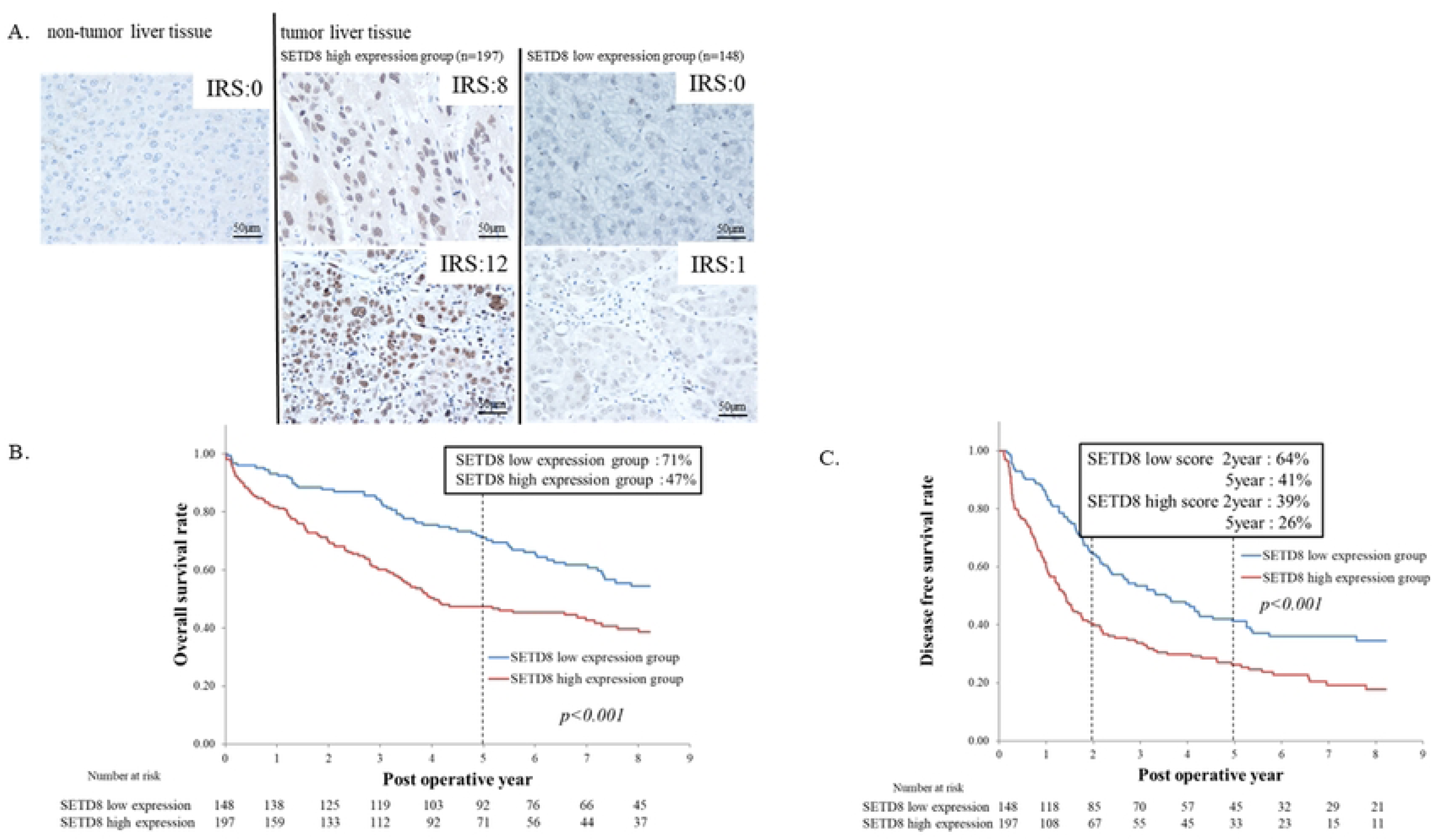
Immunohistochemical analyses and overall survival/disease free survival for HCC. (A) All cases of HCC (n=345) were divided into two groups by immunohistochemical staining pattern of SETD8 depending on IRS. We classified IRS0-5 as SETD8 low expression group (n=148) and IRS6-12 as SETD8 high expression group (n=197). The images represent immunohistochemical staining of SETD8 in tumor and non-tumor liver tissues (x 400 magnification, scale bars; 50µm). (B) SETD8 expression correlated with the five-year overall survival of patients with HCC by Kaplan-Meier method (*p*<0.001). (C) SETD8 expression correlated with the two-year/five-year disease-free survival of patients with HCC by Kaplan-Meier method (*p*<0.001).

**Table 1:**
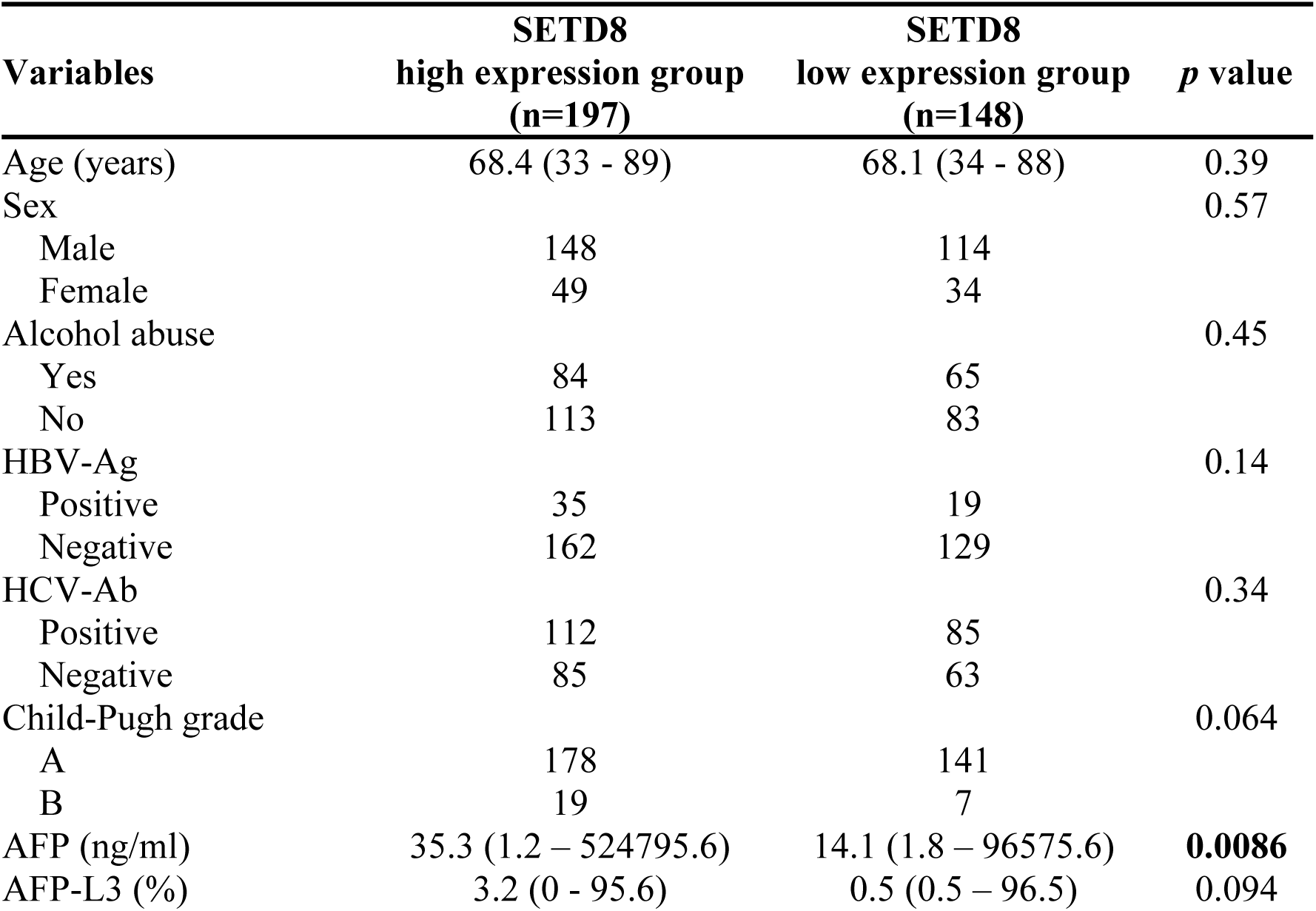
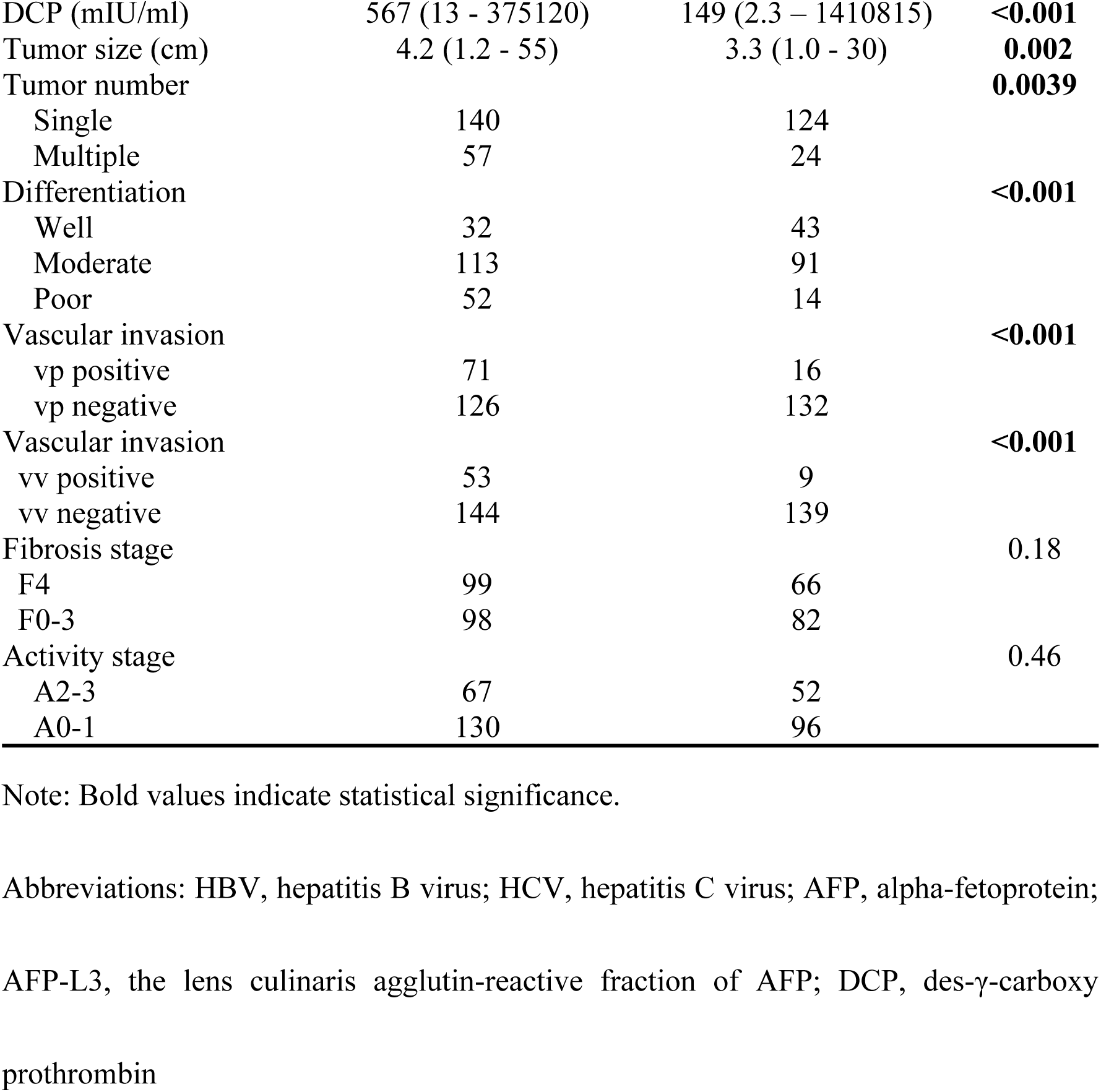
Patients characteristics, clinicopathological findings and preoperative laboratory findings in high and low SETD8 expression groups.

**Table 2.**
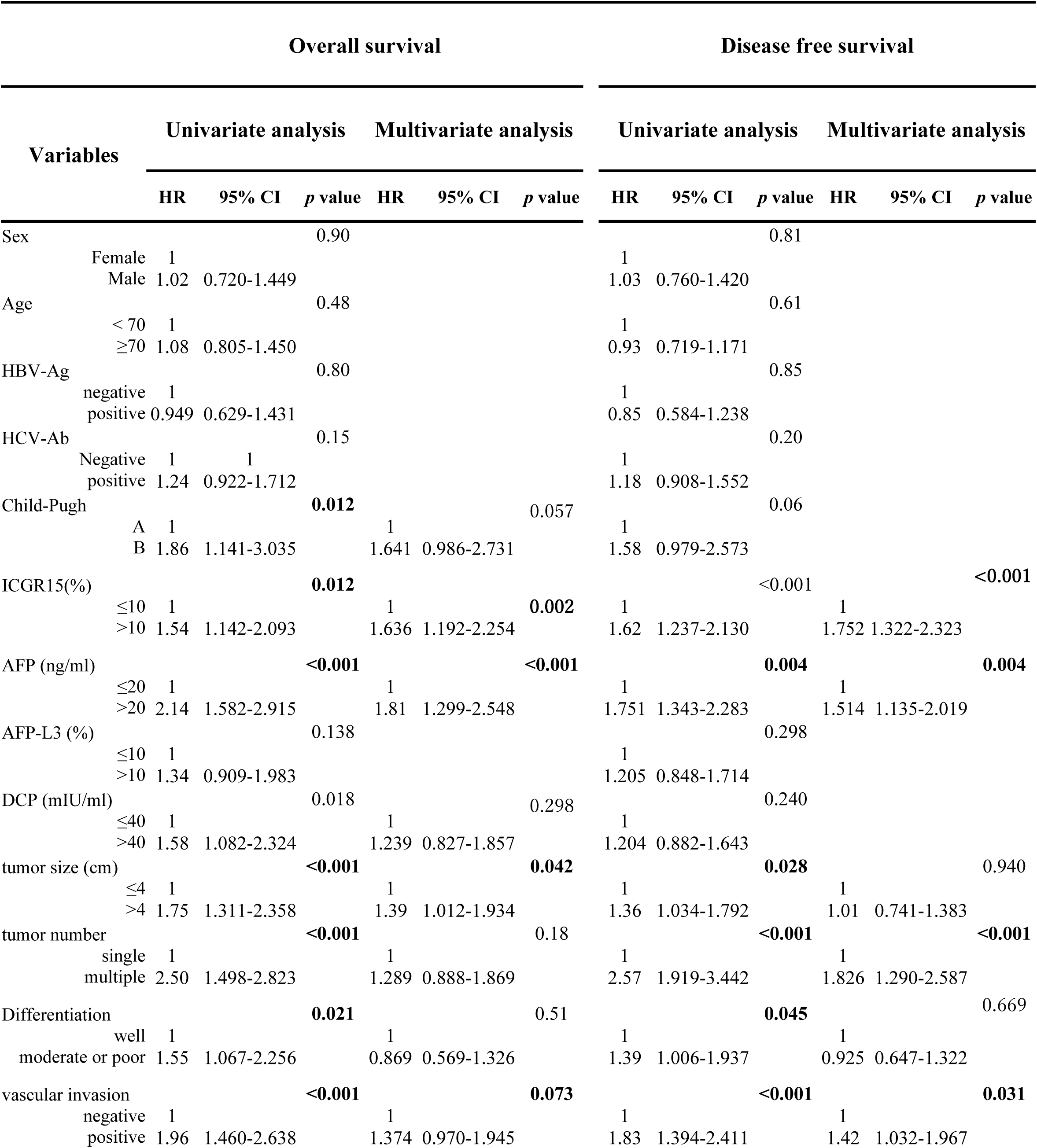
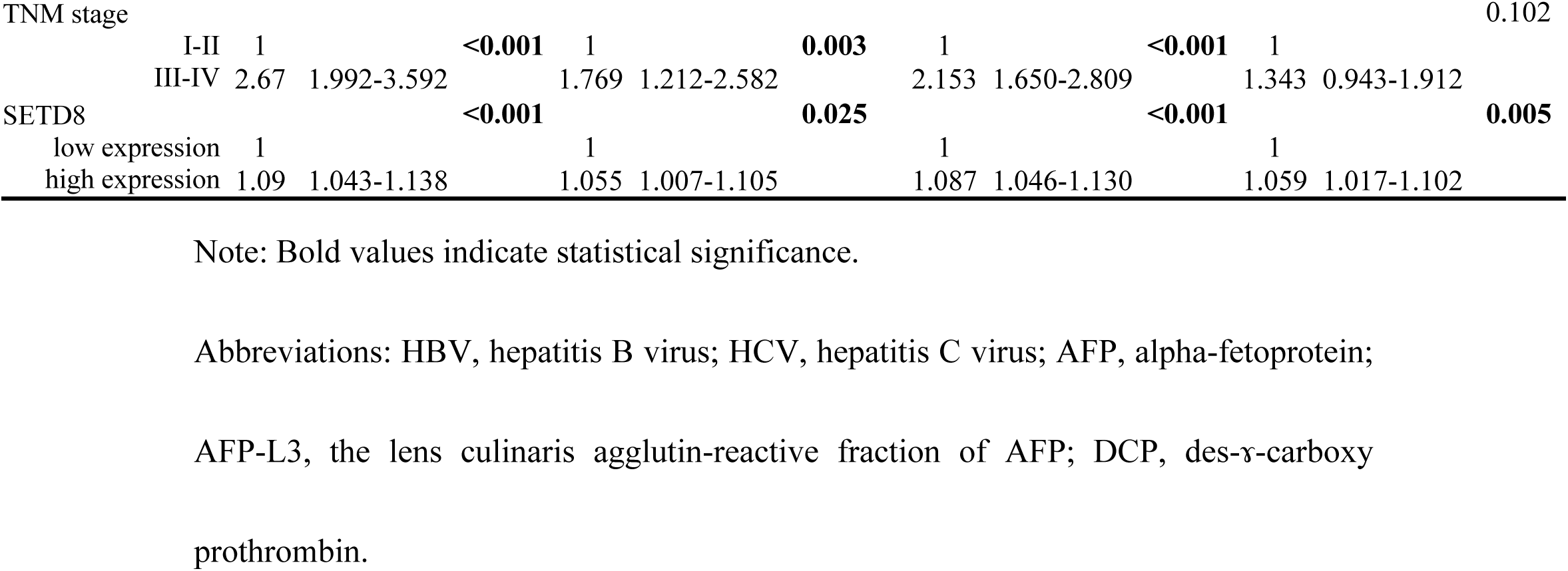
Univariate and multivariate analysis for overall survival and disease-free survival.

### SETD8 regulated HCC cell proliferation, cell cycle, and invasion

Next, loss-of-function analyses were conducted with SETD8-specific siRNA, and siEGFP was used as a negative control. As a result in quantitative real-time PCR, the mRNA expression level of SETD8 in the total RNA extracted from siSETD8-transfected cells had significantly decreased compared with those in the control (Fig 2A and Fig 3A). siRNA functionality has been adequately confirmed, so we then proceeded to perform cell proliferation assay using HCC cell lines transfected with siRNA. HCC cell lines transfected with siSETD8 exhibited a significant inhibition in cell proliferation compared with HCC cell lines transfected with siEGFP (Fig 2B and Fig 3B). Based on these results, we performed cell cycle analysis using flow cytometry to investigate the mechanism of SETD8 in cell proliferation and growth. The proportion of the cell population in each cell cycle phase showed decrease in the S-phase and increase in the G1-phase of siSETD8 compared with the control (Fig 2C, D and Fig 3C, D). SETD8 is suggested to possibly have a role in promoting the transition from the G1 phase to the S phase of the cell cycle. Furthermore, from a pathological perspective, the SETD8 high-expression group exhibited larger tumor sizes, multiple tumors, and more vascular invasion. In the invasion assay, the number of cells that invaded the membrane had significantly decreased in siSETD8-transfected cells compared with siEGFP-transfected cells (Fig 2E and Fig 3E).

**Fig 2.**
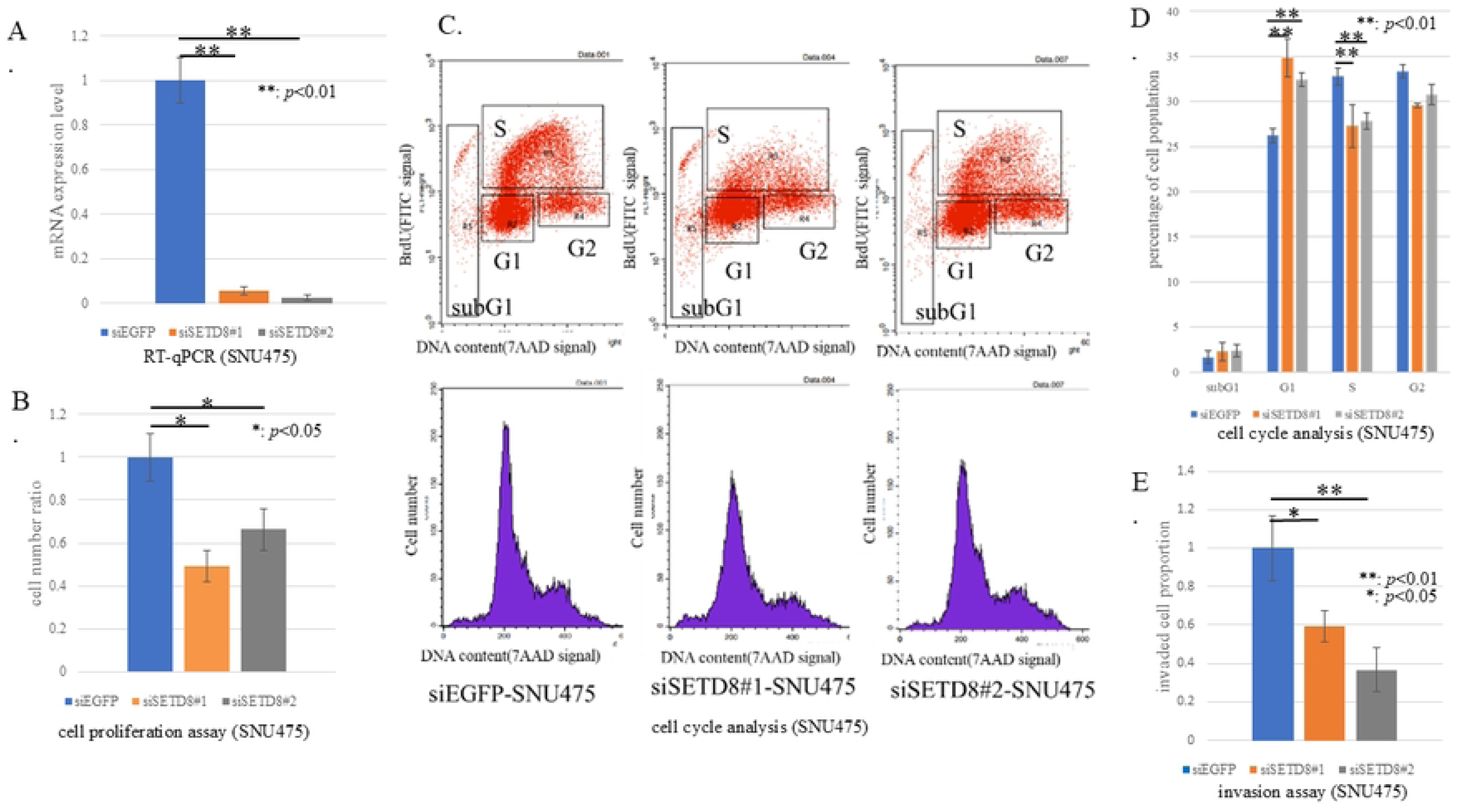
Function analysis of SETD8 using siRNA specific for SETD8 in SNU475. (A) Quantitative real-time PCR showed the suppression of endogenous SETD8 expression of SNU475 using two SETD8 specific siRNAs (#1 and #2). *: *p*<0.05, **: *p*<0.01 (B) SETD8 knockdown could inhibit the growth of SNU475 using cell proliferation assay. (C) Cell cycle analysis after treatment of siEGFP and siSETD8. (D) The percentage of cell number in each phase was counted by flow cytometry. Knocking down SETD8 resulted in an increase in the G1 phase and a decrease in the S phase. **: *p*<0.01 (E) Invasion assay was performed. In HCC cell lines treated with siSETD8, the number of cells that invaded through the basement membrane decreased. *: *p*<0.05, **: *p*<0.01

**Fig 3.**
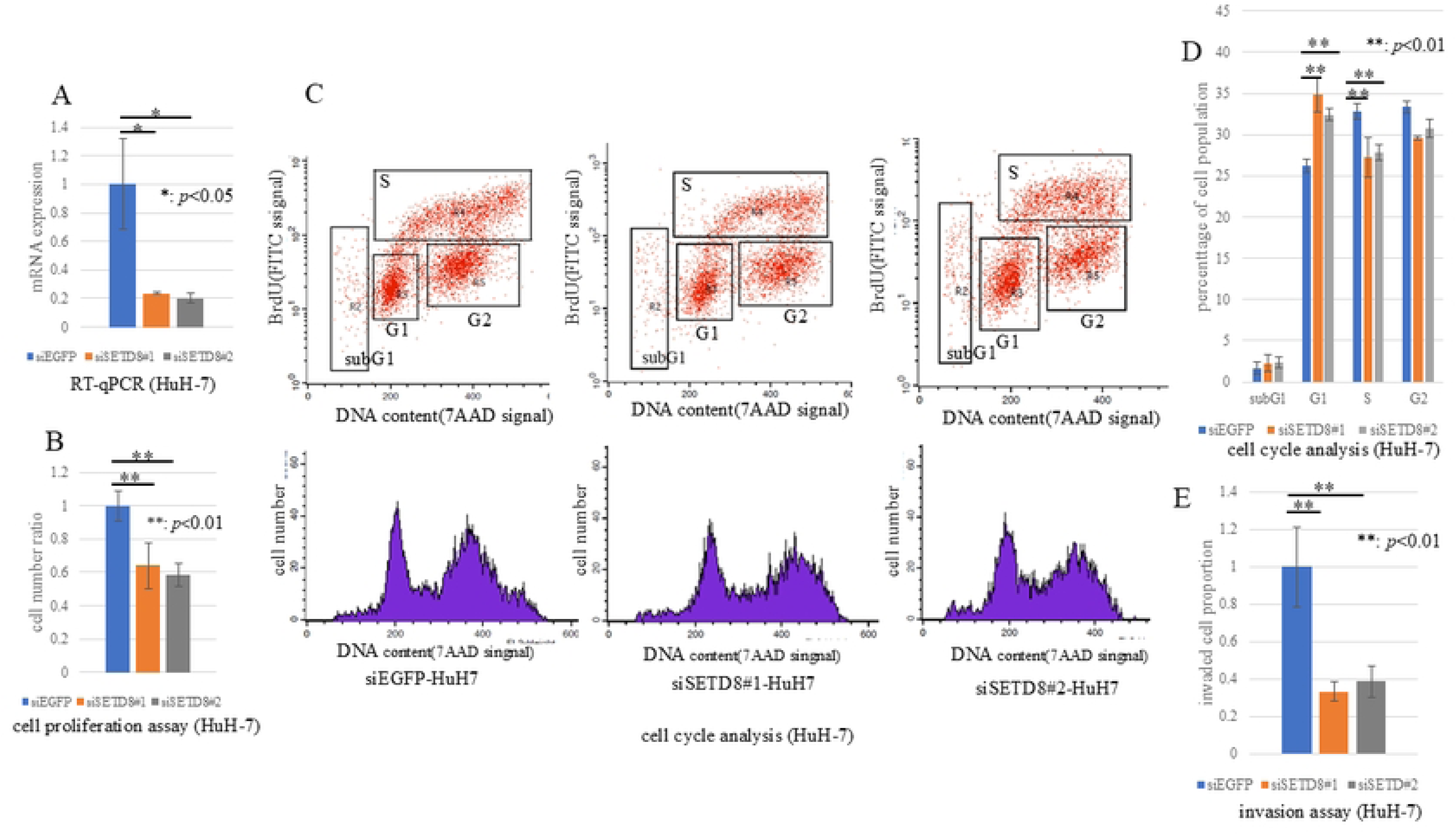
Function analysis of SETD8 using siRNA specific for SETD8 in HuH-7. (A) Quantitative real-time PCR showed the suppression of endogenous SETD8 expression of HuH-7 using two SETD8 specific siRNAs (#1 and #2). *: *p*<0.05 (B) SETD8 knockdown could inhibit the growth of HuH-7 using cell proliferation assay. (C) Cell cycle analysis after treatment of siEGFP and siSETD8. (D) The percentage of cell number in each phase was counted by flow cytometry. Knocking down SETD8 resulted in an increase in the G1 phase and a decrease in the S phase. **: *p*<0.01 (E) Invasion assay was performed. In HCC cell lines treated with siSETD8, the number of cells that invaded through the basement membrane decreased. **: *p*<0.01

### SETD8 was associated with cell growth, proliferation and invasion pathway and regulation of DNA transcription through chromatin remodeling

To investigate downstream pathways associated with SETD8, RNA sequencing analysis was performed using HCC cell line SNU475. Out of the 1388 DEGs, 417 upregulated genes and 971 downregulated genes were identified (see S3 Table). Among the downregulated genes, those related to cancer include cell adhesion-related genes such as carcinoembryonic cell adhesion molecules (CEACAM) 5, CEACAM6 and Kruppel-like factor (KLF) 8, as well as cell cycle-related genes like cyclin D2. Additionally, genes associated with the mitogen-activated protein kinase (MAPK) pathway, such as fibroblast growth factor receptor (FGFR) 2, were also identified. In contrast, among the upregulated genes, those related to cancer include cell adhesion-related genes such as cluster of differentiation (CD) 82, and apoptosis-related genes like Fas (Table 3). We performed gene ontology (GO) analysis based on the extracted DEGs lists (see S4 Table). In GO analysis, GO terms related to cell growth, proliferation, and invasion, such as extracellular signal-regulated kinase (ERK), transforming growth factor beta receptor (TGF-β), FGFR, and phosphatidylinositol 3-kinase (PI3K), were extracted. As a result from GO analysis, we identified clusters related to transcriptional regulation of genes, cancer proliferation, progression and invasion, such as chromatin remodeling, protein kinases, and cell adhesion (Fig 4).

**Fig 4.**
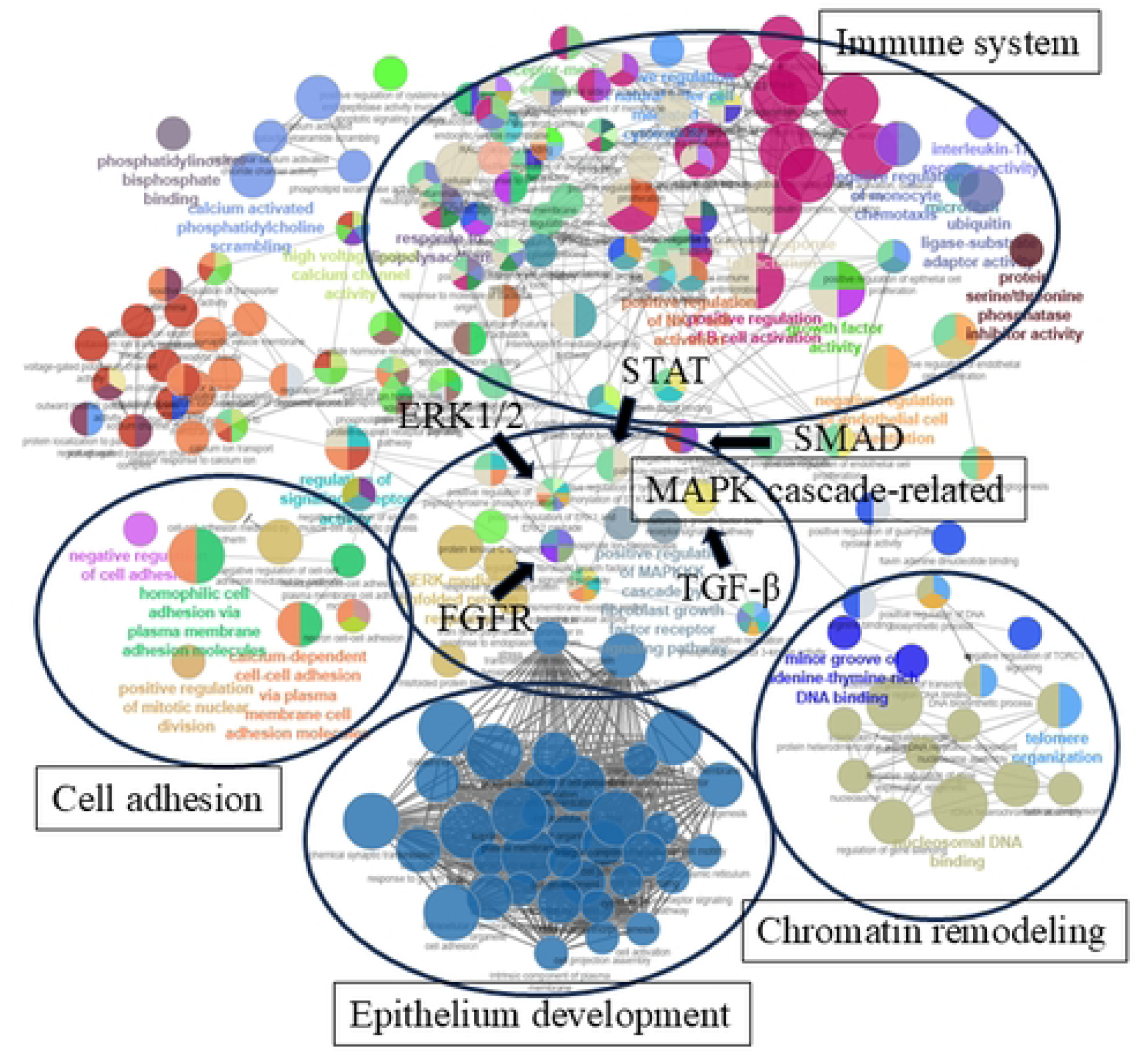
Result of RNA sequence. Pathway analysis associated with SETD8 using gene ontology analysis visualized the results of GO analysis using Cytoscape. Classified into clusters such as ‘cell adhesion’, ‘MAPK cascade-related’, ‘chromatin remodeling’, ‘epithelium development’, and ‘immune system’.

**Table 3.**
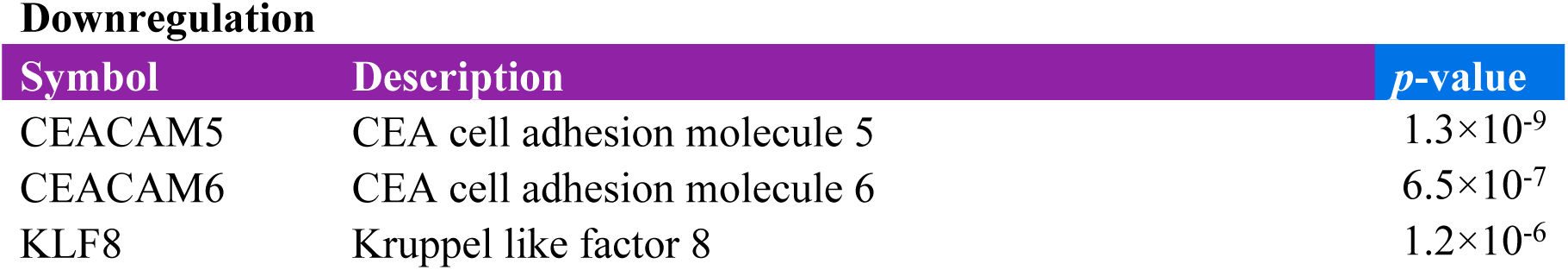
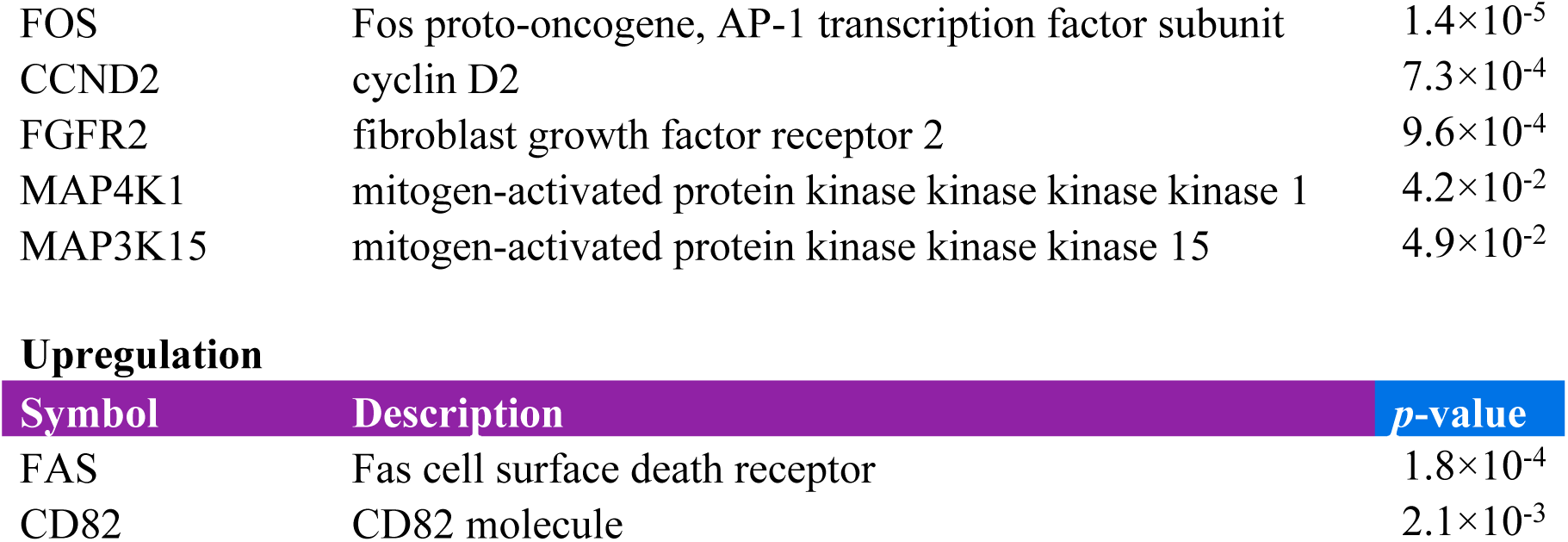
DEG list associated with cancer.

### Tumor growth was inhibited in xenograft model using mice and an HCC cell line in which SETD8 was knocked out via CRISPR/Cas9

Following *in vitro* assay, xenograft experiments were conducted in mice by subcutaneously implanting HuH-7 HCC cell line with SETD8 knockout, which were transfected with CRISPR/Cas9. HuH-7 cells were confirmed to be co-transfected with the SETD8 CRISPR/Cas9 KO plasmid and the SETD8 homology-directed repair (HDR) plasmid incorporating the puromycin resistance gene (Fig 5A). We also observed that the SETD8 mRNA expression levels in the obtained cells were significantly decreased compared with the control (Fig 5B). In control CRISPR/Cas9-treated cells, tumors increased in size over time, whereas SETD8 CRISPR/Cas9-treated cells showed little to no tumor growth (Fig 5C). Only HuH-7 cells were suitable for xenograft experiments, while SNU475 cells failed to establish tumors. To the best of our knowledge, no studies have reported the use of SNU475 cells in a subcutaneous xenograft model.

**Fig 5.**
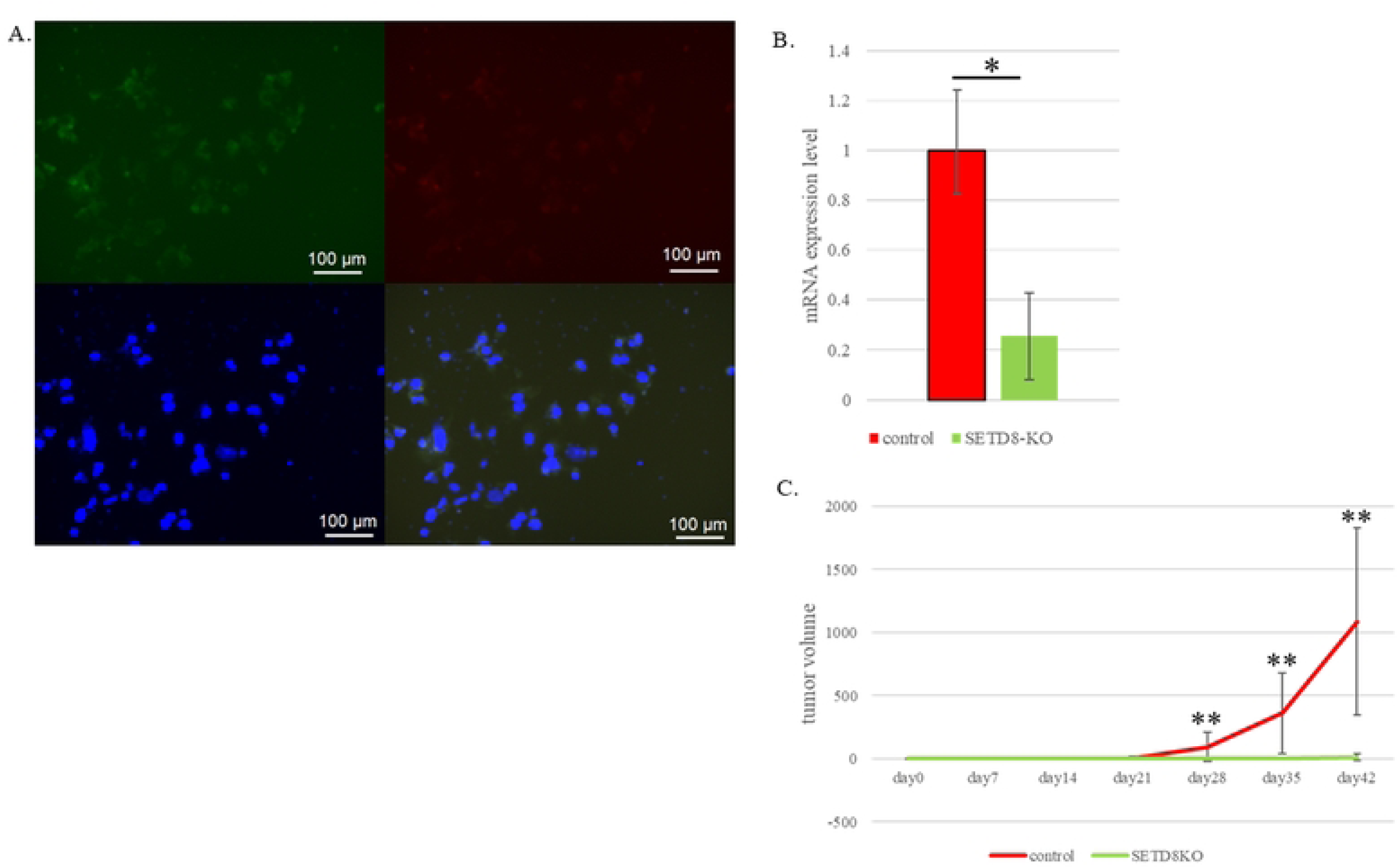
SETD8 KO HuH-7 cells in xenograft mode using CRISPR/Cas9. (A) HCC cell line; HuH-7 with SETD8 knockout were generated using CRISPR/Cas9. Representative images of cell fluorescence immunostaining. Top left: GFP expression (green) in cell transfected with SETD8 knockout plasmid. Top right: Puromycin resistance gene expression (red) in the same cells. Bottom left: DAPI staining (blue) showing cell nuclei. Bottom right: Merged image showing the co-localization of GFP, puromycin resistance gene, DAPI signals. (B) Quantitative real-time PCR showed the suppression of endogenous SETD8 expression of HuH-7 using SETD8 CRISPR/Cas9 KO plasmid. *: *p*<0.05 (C) Subcutaneous xenograft tumors derived from SETD8 knockout HuH-7 cells exhibited little to no growth, whereas control tumors increased over the observation period. **: *p*<0.01

### SETD8 inhibitor UNC0379 suppresses cell proliferation and inhibits cell invasion in HCC cell lines

Preparing future therapeutic targeting of SETD8, functional analyses were performed that employed the SETD8-specific inhibitor UNC0379 as it was thought to be close to clinical application. In similar methodology to siRNA, we conducted cell proliferation assay, cell cycle analysis, and invasion assays with UNC0379. The results showed that in cells treated with UNC0379, compared with the control, cell proliferation was inhibited (Fig 6A), and progression from the G1 phase to the S phase in the cell cycle was suppressed (Fig 6B, C), along with a decrease in cell invasive capabilities (Fig 6D).

**Fig 6.**
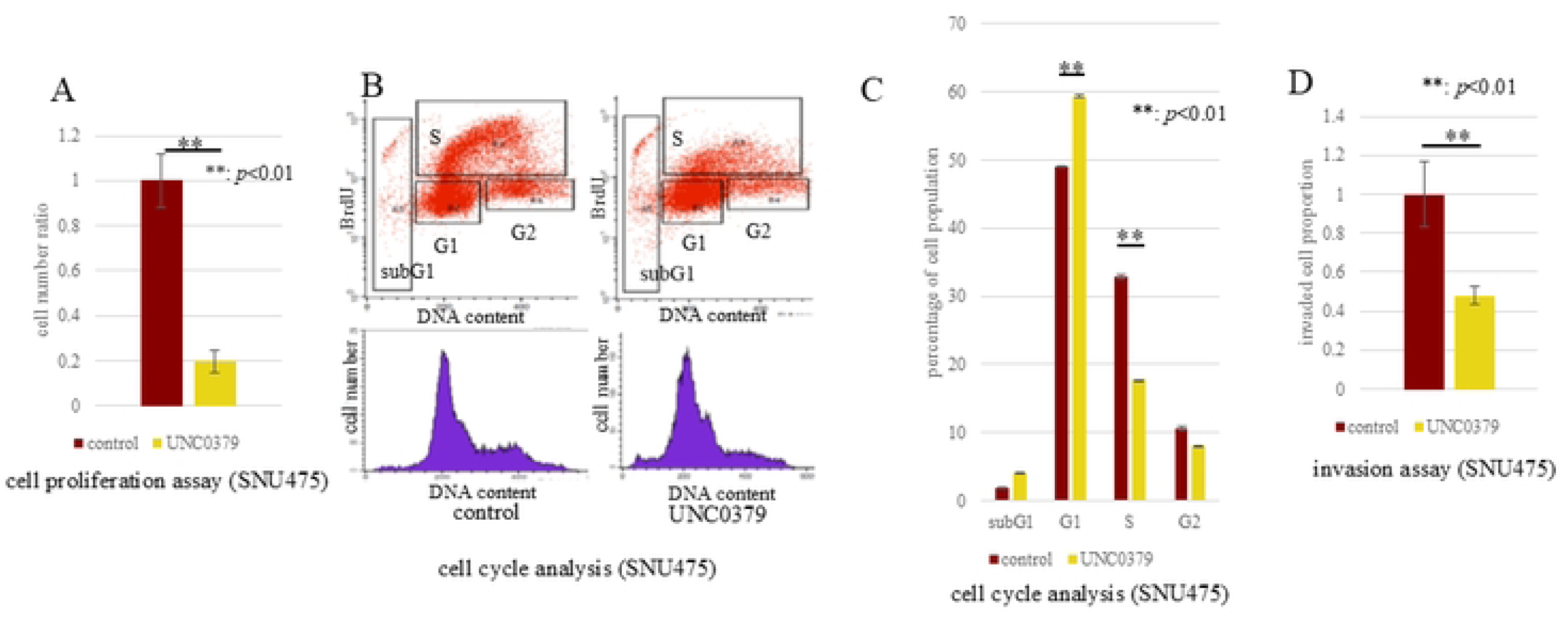
Pharmacological inhibition of SETD8 using UNC0379. (A) The results of cell proliferation assay using SETD8 inhibitor; UNC0379 is presented. In SNU475 treated with UNC0379, cell proliferation is inhibited. **: *p*<0.01 (B) Cell cycle analysis after treatment of UNC0379. (C) In SNU475 treated with UNC0379, inhibition of progression from G1 phase to S phase was observed. **: *p*<0.01 (D) The treatment of SNU475 with UNC0379 led to a reduction in the number of cells invading through the basement membrane. **: *p*<0.01

## Discussion

SETD8 was demonstrated in this study to be a strong prognostic factor for HCC in clinical samples and it played a significant role in the progression of HCC according to various functional analyses. Clinicopathological analyses showed that the SETD8-high expression group exhibited larger tumor sizes and vascular invasion than the low expression group. Furthermore, multivariate analysis revealed that SETD8 is an independent prognostic factor and a risk factor for recurrence in terms of overall survival and disease-free survival. SETD8 is suggested by these results to be significantly associated with the mechanisms of HCC progression. The widespread overexpression of SETD8 across diverse cancer types suggests its potential centrality in cancer progression, underscoring that its importance is not limited to HCC.[21] Notably, the overexpression of SETD8 has been observed in various cancer types, including lung, kidney, and gastric cancers, indicating its importance beyond HCC.[13, 14, 22–27]

Previous research on SETD8 suggested its influence on various biological processes, such as cell cycle regulation, transcription control, and DNA damage response.[12, 15, 28–30] In our study, SETD8 was demonstrated to play a crucial role in the proliferation and invasion of HCC. *In vitro* studies revealed that SETD8 knockdown suppressed cell proliferation, it inhibited progression from G1 phase to S phase in the cell cycle, and it decreased invasion in HCC cell lines. These findings align with the results of our cell proliferation and invasion assays and suggest the clinical importance of SETD8 for HCC.

To further investigate the potential relevance of targeting SETD8, we conducted *in vitro* experiments using the SETD8 inhibitor UNC0379 and *in vivo* experiments using SETD8 KO cell line. The *in vitro* studies demonstrated that treatment with UNC0379 suppressed cell proliferation, inhibited progression from G1 phase to S phase in the cell cycle, and decreased invasion in HCC cell lines. Moreover, *in vivo* studies using xenograft experiments showed that the SETD8 KO cell line exhibited reduced tumor growth compared with the control group. SETD8 inhibitors were suggested to hold promise as a novel approach to cancer treatment. UNC0379 has been shown to inhibit cell proliferation in ovarian cancer, neuroblastoma, glioblastoma, and breast cancer cell lines.[14, 15, 31, 32] Based on these findings, in addition to the multi-kinase inhibitors currently used as key drugs in HCC treatment, experimental results using the SETD8 inhibitor UNC0379 have provided new insights into drug therapy for HCC through the methylation.

Hamamoto et al. proposed the categorization of protein methylations into five different functions: other protein modification (in which methylation subsequently evokes phosphorylation), protein-protein interaction (in which methylation regulates interactions), protein stability (in which methylation inhibits polyubiquitylation), subcellular localization (in which methylation regulates localization between the nucleus and the cytoplasm), and promoter binding (in which methylation promotes transcription factors).[6, 33] The functional analysis of this GO analysis suggests the potential of SETD8-mediated protein methylation to induce phosphorylation. Targeting methylation as a therapeutic approach for HCC holds the promise of enhancing the effectiveness of existing multi-kinase inhibitors, and the co-administration with multi-kinase inhibitors should be a subject of future research. These results suggest that SETD8 could be a promising new therapeutic target and this is perhaps the first step in discovery of anti-cancer drugs related to protein methylation. Tazemetostat, an inhibitor of EZH2, has recently been clinically applied for lymphoma.[34]

The extraction of DEGs in RNA sequencing and GO analysis identified through important biological processes associated with gene sets influenced by SETD8. In GO analysis, several clusters related to cancer were identified. Chromatin remodeling is not limited to methylation but it also encompasses central functions of post-translational histone modifications such as acetylation, phosphorylation, and ubiquitination.[35, 36] The results of GO analysis indicated that SETD8 is involved in transcriptional regulation through chromatin remodeling in HCC. Additionally, kinase-related clusters included pathways such as ERK1/2, FGFR, and TGF-β, which supported the clinical data on tumor size, the results of cell proliferation assay, or cell cycle analysis.[37–39] These findings suggest the possibility that methylation by SETD8 may directly or indirectly influence kinase activity.[25, 40] Clusters related to cell adhesion were also identified, possibly associated with epithelial-mesenchymal transition (EMT), mesenchymal-epithelial transition (MET), or other mechanisms of metastasis.[41] These align with clinical data on vascular invasion and shortened disease-free survival. SETD8 reportedly plays a role in the invasiveness and metastasis of cancer through its interaction with Twist, a crucial regulator of EMT[42], which aligns with and supports the findings of our analysis.

As a limitation of this study, SETD8 has the reported ability of methyltransferase for histone/non-histone protein[12, 29, 30], but we did not investigate the role as an aspect of methyltransferase of SETD8. Further analyses into the methyltransferase activity of SETD8 are required, which may lead to the discovery of a novel target or the pathway of the methylation by SETD8. In addition, this study was a single-center study conducted in Japan belonging to Asian ethnicity, the generalizability of our findings to other ethnic groups or clinical settings may be therefore limited. Further validation in multi-center and international cohorts might be warranted.

In conclusion, this study clarified that SETD8 play a critical role in hepatocellular carcinoma progression through the regulation of cell activity and cell cycle transition. In addition, clinical analysis revealed that overexpressed SETD8 was associated with poor prognosis as an independent prognostic factor.

## Data Availability

All relevant data are within the manuscript and its Supporting Information files.

## Acknowledgement

We acknowledge proofreading and editing by Benjamin Phillis at the Clinical Study Support Center at Wakayama Medical University.

## Declarations

### Ethics Statements

This study was approved by the Wakayama Medical University Hospital Ethics Committee (Approval number: 2521). Animal experiments were approved by the Wakayama Medical University Animal Care and Use Committee (Approval number: 1108).

## Supporting information

**S1 Table: Primer sequence for qRT-PCR**

**S2 Table: siRNA sequences**

**S3 Table: DEGs identified by RNA sequencing.**

This table lists DEGs between siSETD8-treated (sample1-2) and siEGFP-treated (sample2-1) SNU475.

**S4 Table: GO analysis results of DEGs.**

This table lists the GO term identified from the DEGs between siSETD8-treated (sample1-2) and siEGFP-treated (sample2-1) SNU475.

## Notes

### Competing Interest Statement

The authors have declared no competing interest.

### Funding Statement

The author(s) received no specific funding for this work.

### Author Declarations

This retrospective study was approved by the Wakayama Medical University Ethics Committee (approval number: 2521) and conducted in accordance with the principles of the Declaration of Helsinki.

